# The Novel Severe Acute Respiratory Syndrome Coronavirus 2 (SARS-CoV-2) Directly Decimates Human Spleens and Lymph Nodes

**DOI:** 10.1101/2020.03.27.20045427

**Authors:** Zeqing Feng, Bo Diao, Rongshuai Wang, Gang Wang, Chenhui Wang, Yingjun Tan, Liang Liu, Changsong Wang, Ying Liu, Yueping Liu, Zilin Yuan, Liang Ren, Yuzhang Wu, Yongwen Chen

**Author notes:** Corresponding author: **Yongwen Chen** (Ph. D.), Institute of Immunology, PLA, Third Military Medical University, Chongqing, 400038, People’s Republic of China. or **Yuzhang Wu** (Prof. & Chair). Equally to this work (Zeqing Feng, Bo Diao, Rongshuai Wang, Liang Ren, Yuzhang Wu, Yongwen Chen).

## Abstract

While lymphocytopenia is a common characteristic of patients infected by the novel severe acute respiratory syndrome coronavirus 2 (SARS-CoV-2), the mechanisms responsible for this depletion are unclear. Through careful inspection of the spleens and lymph nodes (LNs) from six cases with postmortem examinations, we observed that SARS-CoV-2 could directly infect secondary lymphoid organs to induce cell death. Immunohistochemistry demonstrated ACE2 (angiotensin-converting enzyme 2), the potential receptor of SARS-CoV-2, expresses on tissue-resident CD169^+^ macrophages in spleens and LNs. Immunofluorescent staining confirmed that viral nucleocaspid protein (NP) can be found in ACE2^+^ cells, CD169^+^ macrophages, but not in CD3^+^ T cells or B220^+^ B cells in spleens and LNs. SARS-CoV-2 infection induces severe tissue damage including lymph follicle depletion, splenic nodule atrophy, histiocyte hyperplasia and lymphocyte reductions. Moreover, *in situ* TUNEL staining illustrated that viral infection leads to severe lymphocyte apoptosis, which might be mediated by viral antigens inducing Fas upregulation. Furthermore, SARS-CoV-2 also triggers macrophages to produce IL-6, a proinflammatory cytokine that directly promotes lymphocyte necrosis. Collectively, these results demonstrate that SARS-CoV-2 directly neutralizes human spleens and LNs through infecting tissue-resident CD169^+^ macrophages.

## Introduction

In December 2019, clusters of patients with pneumonia of unknown etiology were reported by the local health facilities in Wuhan, Hubei Province, China.^1,2^ In January 2020, the causative agent of mysterious pneumonia was identified as a novel coronavirus named as 2019 novel coronavirus (2019-nCoV) by the World Health Organization (WHO). This novel coronavirus was renamed as the novel severe acute respiratory syndrome coronavirus 2 (SARS-CoV-2), and the consequent disease was designated as coronavirus disease 2019 (COVID-19).^3,4^ According to the daily report of the WHO, the epidemic of SARS-CoV-2 has so far caused 81961 laboratory confirmed cases and 3293 death in China. Meanwhile, the number of confirmed COVID-19 and fatalities around the world are 465315 and 21031, respectively, by 26^th^ March 2020.^5^

The importance of the secondary lymphoid organs, including spleen and lymph nodes (LNs), for resistance against infection is well established. Tissue-resident macrophages positioned in the splenic marginal zone (MZ) that are among the first cell types to encounter invading pathogens.^6,7^ Similarly, the resident macrophages of the subcapsular sinus and hilar lymph nodes have been shown to play a protective role against viral infections by capturing viral particles.^8,9^ These macrophages present viral antigens to activate specific T cells, to induce activation, clonal proliferation, and subsequent killing of infected target cells through secretion of perforin, granzyme B, and interferon-γ etc.^10-12^ Moreover, these activated T cells also egress from spleen and LNs into blood circulation and play immune surveillance roles.^13^ Unfortunately, our previous work has shown that lymphocytopenia is prevalent in COVID-19 patients, especially in aged and critically ill cases, suggesting that the immune system from COVID-19 patients might be neutralized by infection.^14^ Therefore, understanding the destinies of lymphocytes in secondary lymphoid organs is critical for understanding SARS-CoV-2 viral infection, and the identification of methods to boost lymphocytes and enhance the host immunity *in vivo*.

In this report, spleens and LNs from six COVID-19 patients with postmortem examinations were collected. The virions in LNs were visually observed by transmission electronic microscope (EM) and pathological damage was analyzed by hematoxylin and eosin (H&E) staining. Moreover, viral nucleocapsid protein (NP) antigen, cell apoptosis and proinflammatory cytokine expression were measured by immunohistochemistry.

## Methods

### Patients

Postmortem autopsies were conducted on six COVID-19 patients who had been admitted in Jinyintan Hospital in Wuhan, Hubei province, China. Diagnosis of COVID-19 was based on the New Coronavirus Pneumonia Prevention and Control Program (5^th^ edition) published by the National Health Commission of China. All the patients were laboratory-confirmed positives for SARS-CoV-2 by use of quantitative RT-PCR (qRT-PCR) on throat swab samples. Moreover, three cases of spleens and LNs from normal healthy controls who were death due to traffic accident were also involved. This study was approved by the National Health Commission of China and Ethics Commission of General Hospital of Central Theatre Command ([2020]017-1) and Jinyintan Hospital (KY-2020-15.01). Written informed consent was waived by the Ethics Commission of the designated hospital for emerging infectious diseases.

### Tissue morphology detection

Due to the special infection-control precaution of handling deceased subjects with COVID-19, postmortem examination was performed in a designated pathology laboratory. The spleens and hilar lymph nodes from 6 cases were collected by standard examination at autopsy. The histopathology and pathological characteristics were analyzed by H&E staining. Briefly, paraffin-embedded tissue blocks were cut into 3 μm slices and mounted onto poly-lysine-coated glass slides, tissues were incubated with hematoxylin for 5 minutes, after 1 min of dehydration by 100% alcohol, section were further treated with eosin for 30 seconds. Sections were mounted and the results were viewed using a light microscope (Zeiss Axioplan 2, Berlin, Germany).

### Immunohistochemistry

The protocol used for immunohistochemistry was performed as published previously.^15^ Briefly, paraffin-embedded tissue blocks were cut into 2∼3μm sections and mounted on poly-L-lysine-charged glass slides. Sections were de-waxed and rehydrated, and antigen retrieval was performed by microwaving in 10 mM citrate buffer (pHL6.0). Endogenous peroxidase activity was blocked by incubation with a solution of 0.5% hydrogen peroxidase (H_2_O_2_) in 50 % methanol for 1 h. The sections were then incubated in 3% BSA plus 0.1% Nonidet P-40 in PBS for 1 h at RT to block nonspecific binding. Sections were then incubated overnight at 4 °C with primary antibodies that had been diluted in 1 % BSA. These antibodies are including anti-SARS-CoV-2 nucleocaspid protein (NP) antibodies (clone ID: 019, 1:100, rabbit IgG; Sino Biological, Beijing), anti-ACE2 (clone ID: 10108-RP01, 1:100, rabbit IgG; Sino Biological), anti-CD68 (Clone ID:KP1, 1:100, mouse IgG1; BIO-RAD), anti-CD169 (Clone ID:7-239, 1:100, mouse IgG1; Biolegend), anti-B220 (Clone ID:123C3, 1:100, mouse IgG1; BIO-RAD), anti-Fas (ID:48095942, 1:100, mouse IgG; Thermofisher), anti-FasL (sc-834, 1:100, rabbit IgG; Santa Cruz), anti-IL-6 (sc-130326, 1:100, mouse IgG2b; Santa Cruz) or rabbit-isotype antibody controls (1:100; Dako). After washing, the sections were incubated with the corresponding secondary antibodies for 1 h at RT. The Vecta-stain ABC kit (Vector Laboratories, San Diego, CA, USA) was used for the avidin-biotin complex method according the manufacturer’s instructions. Sections incubated with isotype-matched, concentration-matched immunoglobulin without primary antibodies were used as isotype controls. Peroxidase activity was visualized with the DAB Elite kit (K3465, DAKO, Copenhagen, Denmark), and brown coloration of tissues represented positive staining. The sections were lightly counterstained with hematoxylin, dehydrated through an ethanol series to xylene and mounted. Finally, sample sections were viewed using a light microscope (Zeiss Axioplan 2).

### Immunofluorescence double-staining

For immunofluorescence double-staining, the sections were incubated with primary anti-NP, anti-CD68, anti-CD169, anti-B220, anti-IL-6 antibodies at 4 °C overnight. After washing with PBS (3 washes, 5 min per wash), the sections were incubated with Alexa Fluor^®^ 555-conjugated goat anti-mouse/rabbit IgG antibodies (Invitrogen, San Diego, CA, USA) or Alexa Fluor^®^ 488-conjugated goat anti-mouse/rabbit IgG1 antibodies (Invitrogen) for an 1 h. Finally, the sections were incubated with 1 μg/ml DAPI (Sigma, St. Louis, MO, USA) for 10 min to stain the nuclei. Sections incubated with the appropriate isotype control primary antibodies and fluorescently labeled secondary antibodies were used as negative controls. The results were analyzed using fluorescence microscopy (Zeiss Axioplan 2).

### TUNEL staining

Cellular apoptosis was measured by TUNEL staining according to the manufacturer’s instructions (Roche, Berlin, Germany). The proportion of TUNEL-positive nuclei in spleens and LNs was determined through image analysis of the histological sections. Photomicrographs were captured and analyzed using Image Pro-Plus 5.0 software (Media Cybernetics, Silver Spring, MD).

### Statistical analysis

Statistical analyses were performed using GraphPad Prism version 8.0 (GraphPad Software, Inc., San Diego, CA, USA). Qualitative variables were expressed as numbers. p values are from t test.

## Results

### 1. ACE2 expresses on macrophages in spleens and LNs

Immunohistochemistry was used to detect the expression of ACE2, the potential receptor for SARS-CoV-2, in 6 cases of spleens and hilar LNs from 6 autopsies as well as normal human tissues. Results showed that ACE2^+^ cells were observed in spleens and LNs from normal healthy and autopsies. In LNs, the expression of ACE2 is restricted on cells within medulla (**Figure 1 A** and **C**), whereas ACE2^+^ cells are found in red pulp of spleens (**Figure 1 B** and **D**), and tissues from autopsies have slightly higher levels of ACE2^+^ cells than normal controls. Here ACE2 expression in normal kidneys were used as positive controls (**Figure 1E**), and sections without primary antibodies were used as isotype controls (**Figure 1F**), demonstrating ACE2^+^ cells are positions in human spleens and LNs.

**Figure 1.**
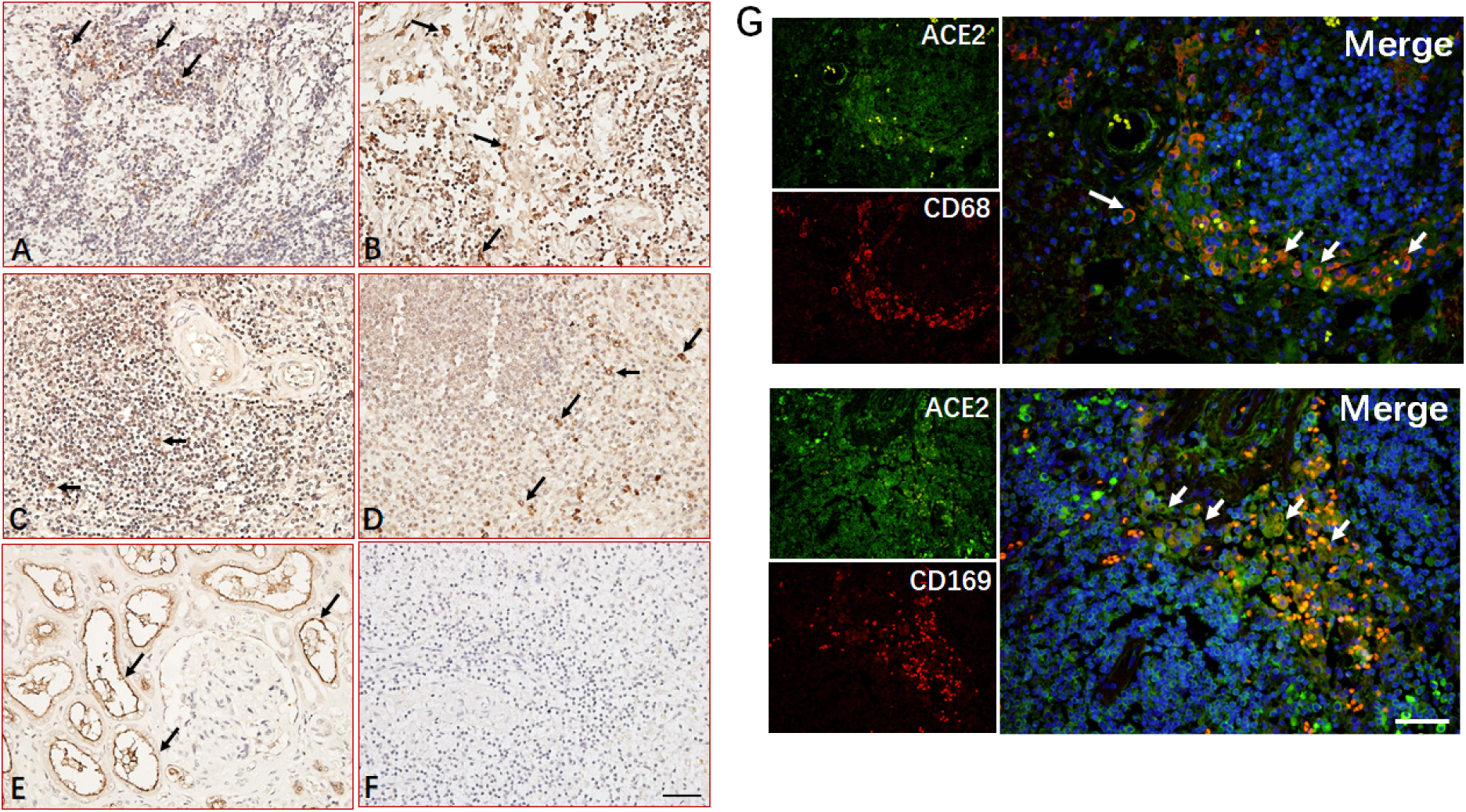
Immunohistochemistry analyzed ACE2 antigen in spleen and LN tissues. The expression of ACE2 was detected in sections from LN (**A**) and spleen (**B**) from normal healthy as well as sections from LN (**C**) and spleen (**D**) of COVID-19 patients undergoing postmortem examination. (**E**) Sections from normal human kidneys are used as positive control. (**F**) Antibodies isotype control. **(G**) Immunofluorescent double staining analyzed ACE2 express on CD68 and CD169 positive macrophages. Arrow indicated positive cells. Scale bar= 100 μM.

We next examined what kinds of cells are positive for ACE2, and immunofluorescent double staining showed that ACE2 expresses on CD68^+^ macrophages in LNs (**Figure 1G**). CD169^+^ macrophages are a subpopulation of tissue-resident macrophages positioned in the splenic marginal zone (MZ) where they encounter invading pathogens.^6,7^ In parallel, CD169^+^ macrophages also reside in the subcapsular sinus of LNs and have been shown to capture viral particles from draining LNs.^8,16^ Immunofluorescent double staining further illustrated that the ACE2 antigen is observed in CD169^+^ macrophages in the subcapsular sinus of LNs (**Figure 1G**). Collectively, these results demonstrated that ACE2 expresses on macrophages in spleens and LNs.

### 2. SARS-CoV-2 directly infects macrophages in spleens and LNs

We next investigated the expression of viral NP antigen in secondary lymphoid tissues.

Immunohistochemistry showed that SARS-CoV-2 NP antigens could be seen in spleens and LNs from all of these six autopsies, while being absent in sections from normal healthy controls. In the spleen, viral NP^+^ cells were primarily distributed in red pulp and blood vessels, although positive cells were also occasionally observed in white pulp. In lymph nodes, the NP^+^ cells were seen in cells within marginal sinus of lymph nodules as well as capillaries, some positive cells are also found in germinal centers, and NP antigen was observed in cytoplasm, whereas nucleus is negative for NP expression **(Figure 2A)**.

**Figure 2.**
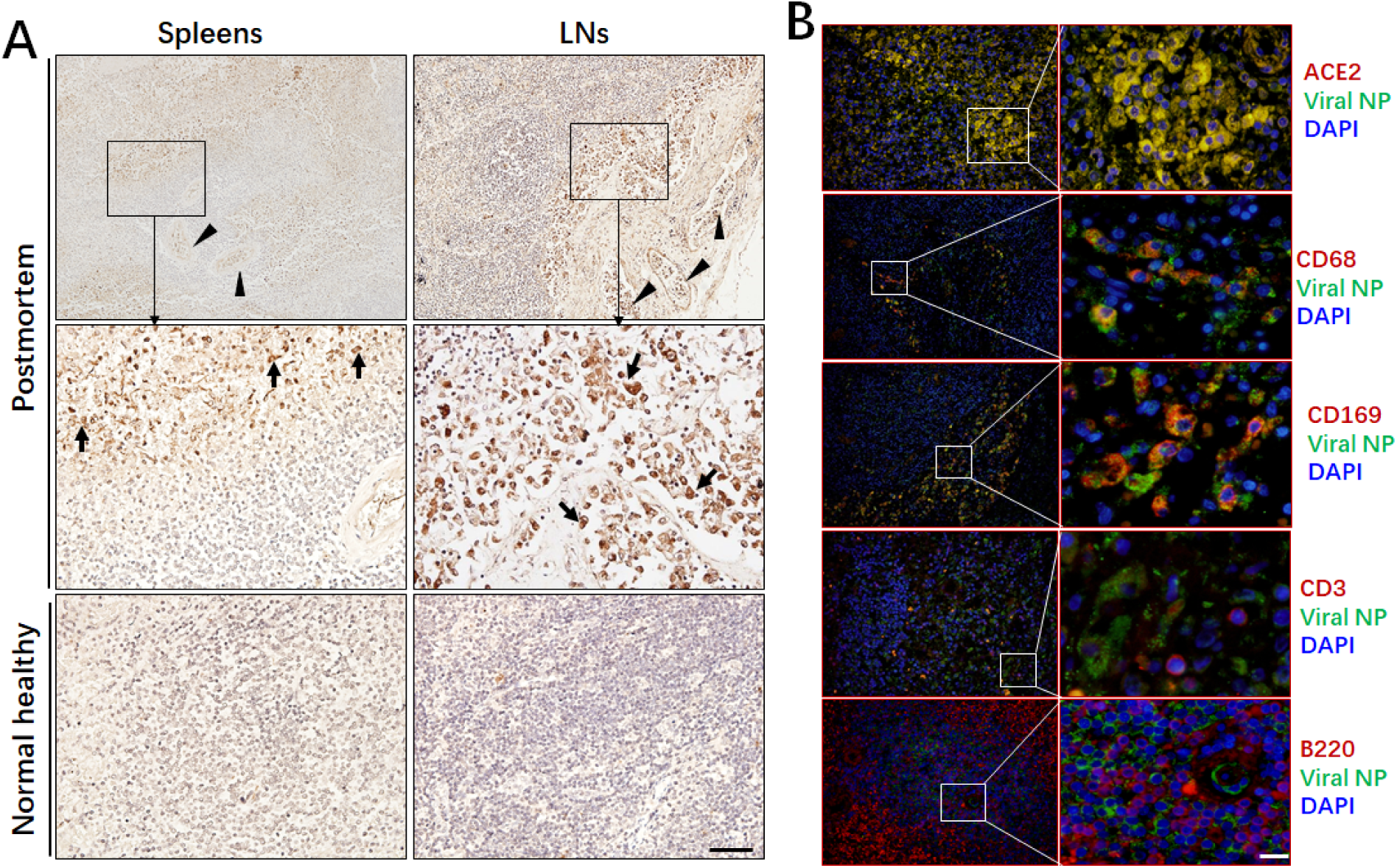
Immunohistochemistry analyzed SARS-CoV-2 NP antigen in spleen and LN tissues. **(A)** The expression of viral NP antigen was detected by immunohistochemistry in spleen and LN tissues from COVID-19 patients undergoing postmortem examination and age-match normal healthy controls. **(B**) Immunofluorescent double staining analyzed viral NP antigen expression. Arrow indicated viral NP positive cells. Scale bar= 100 μM.

We then performed addition staining to identify the LN cell types that are positive for viral NP antigen. Immunofluorescent double staining confirmed that the majority of NP antigen is found in ACE2^+^ cells and CD68^+^ macrophages, including cells in capillaries, while, neither CD3^+^ T cells nor B220^+^ B cells are positive for NP protein **(Figure 2B)**. Moreover, immunofluorescent double staining also further illustrated that the viral NP antigen is observed in CD169^+^ macrophages in the subcapsular sinus of LNs (**Figure 2B**). Collectively, these results suggest SARS-CoV-2 infects macrophages, especially resident CD169^+^ macrophages in spleen and LNs.

### 3. SARS-CoV-2 destroys human spleen and LNs

Since SARS-CoV-2 is known to be highly cytotoxic, its persistence in secondary lymphoid organs may well induce lymphocytopenia. To confirm this possibility, 6 hilar lymph nodes of 1.0∼ 1.5 cm in diameter from 6 autopsies were collected and histopathological examination was performed by H&E staining. In contrast to normal healthy controls, the lymph follicles and paracortical areas in viruses-infected tissues are not identifiable, with necrotic and apoptotic lymphocytes being widely distributed, causing a significant reduction of total lymphocytes, including cells in T and B zones. Moreover, interstitial blood vessels had proliferated, expanded and were congested (**Figure 3**). Similarly, the total lymphocyte counts are significantly lower in sections from viruses infected spleens, which were also dominated by lymphocytes undergoing necrosis and apoptotic. Moreover, the spleens were congested, hemorrhagic, and lacking lymphoid follicles. Additionally, the spleen corpuscles are atrophic, with clear interstitial vessels and fibrous tissue hyperplasia in the splenic sinus (**Figure 3**). These results demonstrate that SARS-CoV-2 causes severe damage in human spleen and LNs.

**Figure 3.**
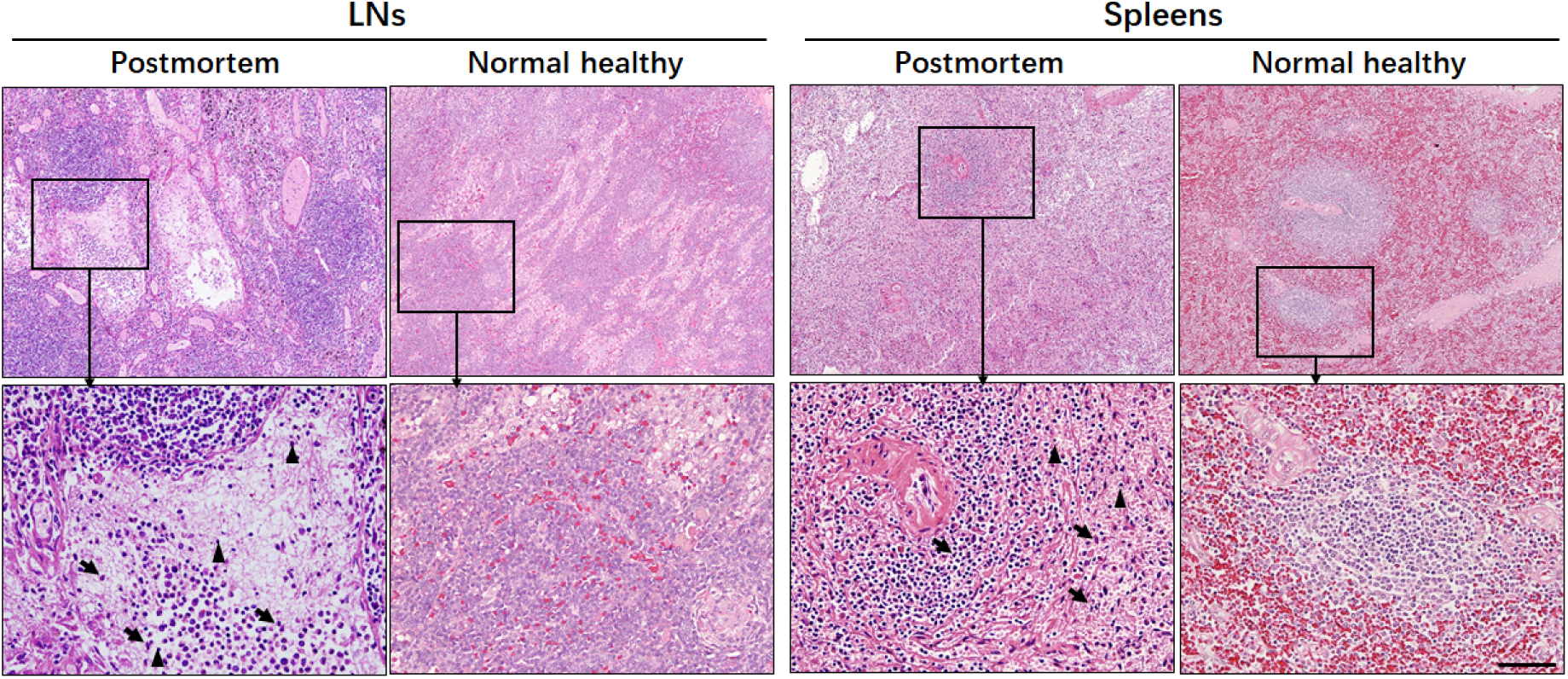
Representative H&E staining of spleen and LN tissues from one COVID-19 patient undergoing postmortem examination and age-match normal healthy control. Sections were stained by H&E, arrow indicated apoptotic lymphocytes; arrow head indicated necrosis cells. Scale bar= 100 μM.

### 4. SARS-CoV-2 induces lymphocyte apoptosis *via* enhancing Fas signaling

*In situ* TUNEL staining showed that viral infected spleen and LNs manifested strong lymphocyte apoptosis, whereas, apoptotic cells in tissues from age matched normal healthy controls are undetectable (**Figure 4A, B**), suggesting SARS-CoV-2 induces cell apoptosis.

**Figure 4.**
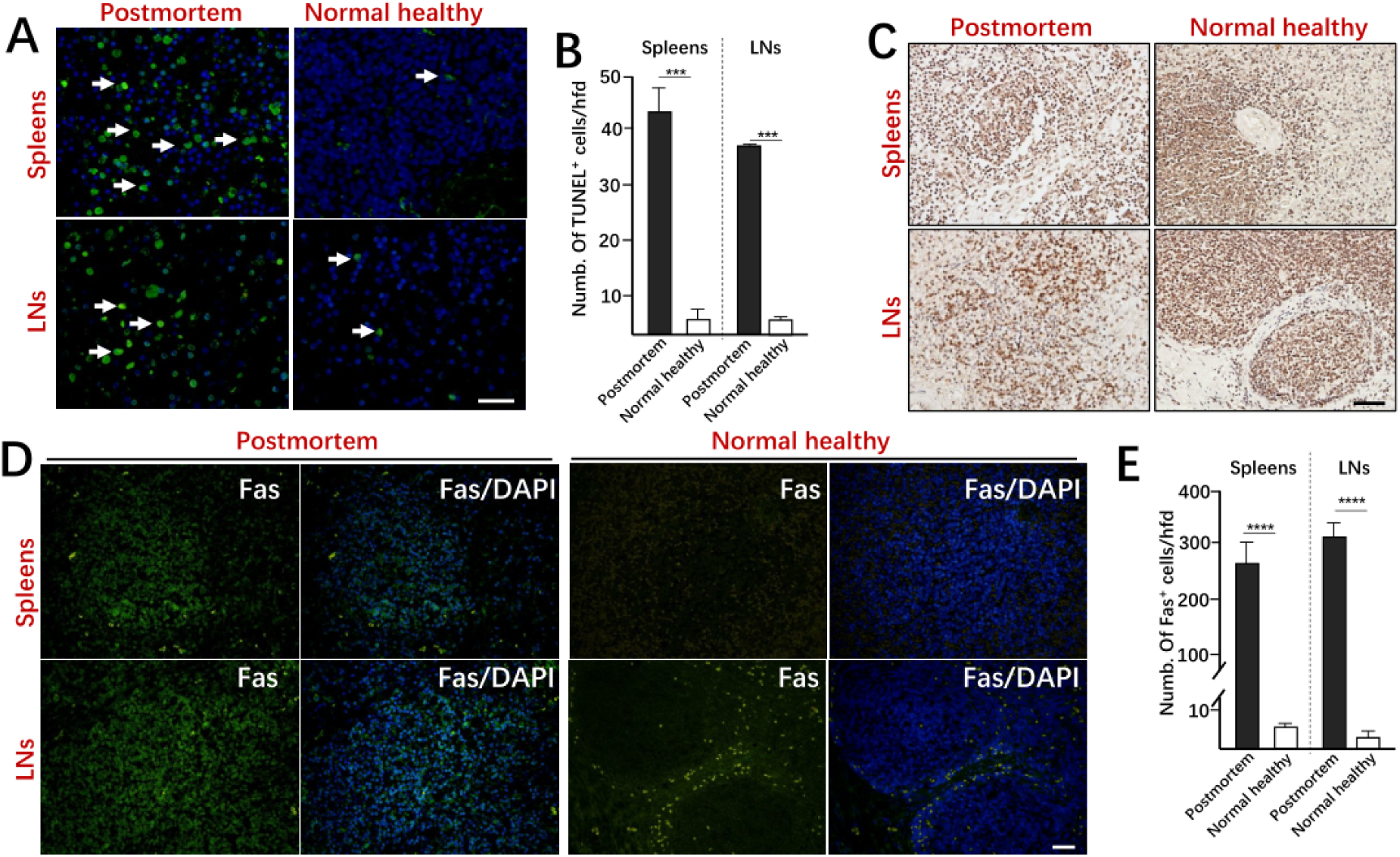
SARS-CoV-2 induces lymphocyte apoptosis *via* enhancing Fas signaling. **(A)** Cell apoptosis was detected by *in situ* TUNEL staining, (**B**) statistical analysis of apoptotic lymphocytes in sections from postmortem and normal healthy controls. (**C**) Immunohistochemistry detected FasL expression. (**D**) Immunofluorescent staining analyzed Fas expression. (**E**) Statistical analysis of Fas^+^ cells in sections from postmortem and normal healthy controls. Scale bar= 100 μM, \*\*\**p*<0.001, \*\*\*\**p*<0.00001.

Because SARS-CoV-2 virus does not infect lymphocyte directly, we think the persistence of viral antigen could constitutively activate T cells and B cells in spleen and LNs, thus causing activation-induced cell death (AICD), which is mediated by Fas/FasL signaling.^17-19^ Immunohistochemistry showed that spleen and LN tissues from autopsies and normal healthy controls manifested similar levels of FasL expression (**Figure 4C**). On the other hand, the expression of Fas was significantly upregulated in virus infected spleens and LNs, whereas, the expression of Fas is absence in sections from normal healthy controls, as detected by immunofluorescent staining (**Figure 4D, E**). These results suggest SARS-CoV-2 promotes lymphocyte apoptosis through enhancing Fas expression.

### 5. SARS-CoV-2 triggers macrophages to secret IL-6 and promote lymphocytopenia

The dysregulation of the proinflammatory cytokines like TNF-α and IL-6 has been described to promote cell necrosis and apoptosis.^20,21^ We also reported that IL-6 might involve in mediating lymphocyte reduction in severe COVID-19 patients.^14^ Immunohistochemistry showed that high levels of IL-6 rather TNF-α were observed in virus infected spleen and LNs compared with sections from normal healthy controls (**Figure 5A**). Moreover, immunofluorescent double staining showed that IL-6 is primarily produced by infected macrophages (**Figure 5B**). Increased transcription of several cytokines in macrophages has been described to be activated by SARS-Spike protein.^22,23^ Since SARS-CoV-2 and SARS share a highly similar Spike protein based on amino acid sequence, we reasoned that a similar phenomenon might occur with SARS-CoV-2 Spike. We thus treated human macrophage line-THP1 cells with Spike protein *in vitro*, and found that SARS-CoV-2 Spike protein can trigger increased *Il6* expression independently of changes in *Il1b* and *Tnfa* gene transcription (**Figure 5C**). Collectively, these results suggest that SARS-CoV-2 viruses and the Spike protein, can trigger macrophages to secrete IL-6, which may accelerate lymphocytopenia.

**Figure 5.**
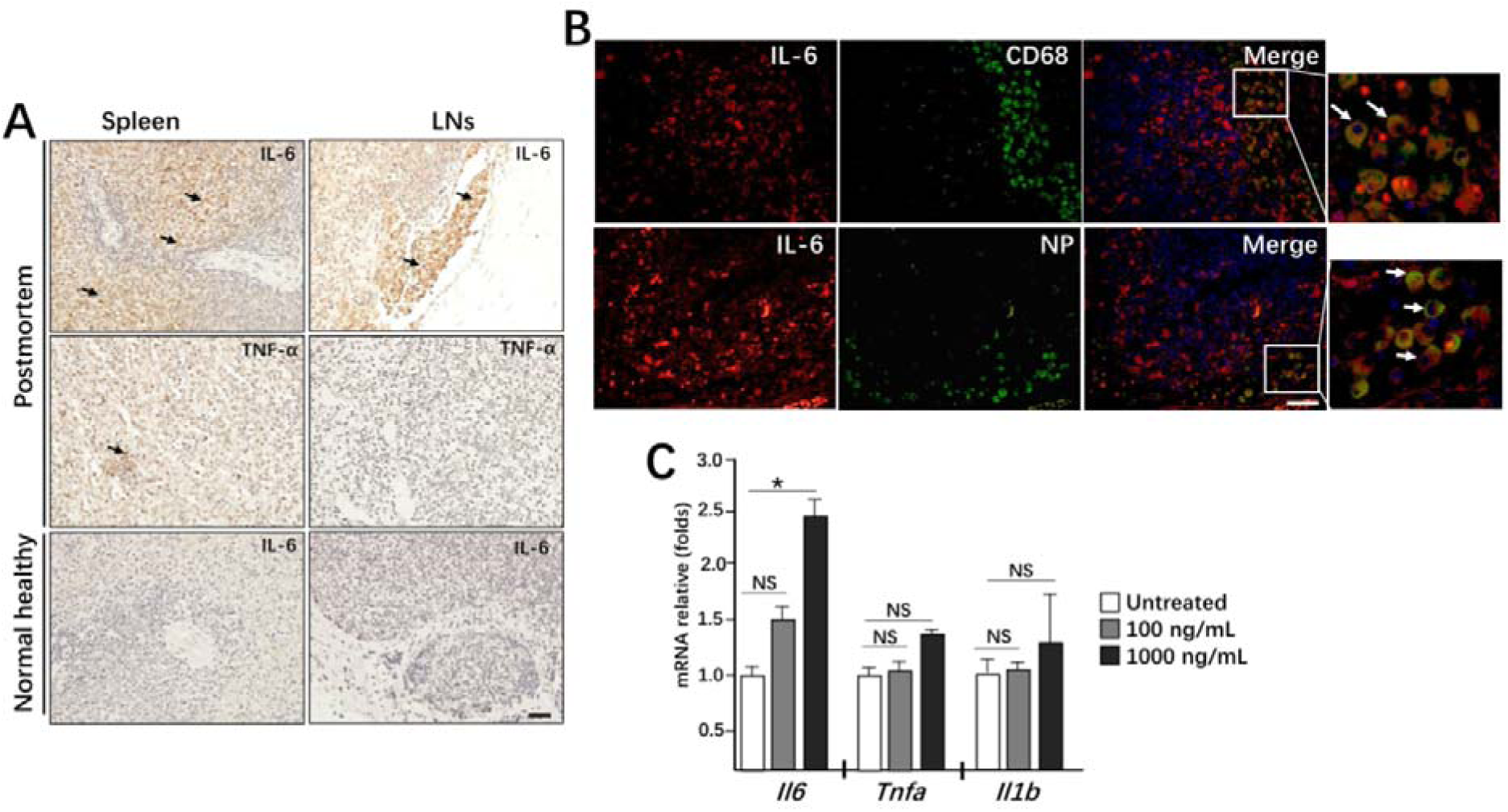
SARS-CoV-2 triggers macrophages to secret IL-6 and promote lymphocytopenia. **(A)** The expression of IL-6 and TNF-α was detected by immunohistochemistry in spleen and LN tissues from COVID-19 patients undergoing postmortem examination and age-match normal healthy controls. **(B**) Immunofluorescent double staining analyzed IL-6 expression. Arrow indicated viral positive cells. Scale bar= 100 μM. (C) Human macrophage line-THP1 cells were stimulated with Spike protein *in vitro*, the transcription of indicated genes after 12h. NS: not significant different, *\*p*<*0*.*05*.

## Discussion

The COVID-19 is an acute infectious disease that caused by SARS-CoV-2 infection. It has been a worldwide epidemic outbreak because SARS-CoV-2 is highly infectious and does not respond to conventional antiviral treatment. Due to global research and collaboration, important progress has been made in understanding its aetiology and epidemiology. Many studies have reported that SARS-CoV-2 infection might induce immune damage, and the reduction of lymphocytes is very common in severe or aged COVID-19 patients.^14,24-26^ However, no data are available regarding whether this virus can directly infect human secondary lymphoid organs to cause lymphocytopenia. We here showed that the expression of SARS-CoV-2 receptor ACE2 is found in macrophages in human spleen and hilar LNs from autopsy as well as normal healthy (**Figure 1**). SARS-CoV-2 is a cytopathic virus which can directly induce cellular damage of target cells during infection, and H&E staining showed that acute tissue damage and lymphocyte reduction occurred in all detected LNs and spleens as anticipated. In particular, the hilar LNs were congested and hemorrhagic, with depletion of lymphocytes and accompanied by subcapsular sinus histiocytosis. Some cases also showed enlargement of abdominal lymph nodes, which have reduced number of germinal centers. Similarly, all of the spleens exhibited atrophy of white pulps, with complete loss in some cases. The red pulp is markedly congested and hemorrhagic (**Figure 3)**. This phenomenon is consistent with older reports of patients infected with SARS-CoV.^27,28^ Taken together, these results demonstrate that SARS-CoV-2 directly infects LNs and spleens to cause severe tissue damage.

We then sought to investigate the route and identities of the cells that could potentially contribute to SARS-CoV-2 transports into spleens and LNs. From our *in situ* immunohistochemistry, we showed that viral NP antigen was restricted to the macrophages of the infected tissues, while, human T cells and B cells were negative for viral NP antigen, suggesting SARS-CoV-2 cannot directly infect T and B cells (**Figure 2B**). Most importantly, viral NP^+^CD68^+^ macrophages were observed in capillaries of spleens and LNs, indicating that SARS-CoV-2 migrates into spleens and LNs through macrophages. An interesting finding is that overwhelming numbers of NP^+^ cells are observed in red pulp of spleens and within marginal sinus of lymph nodes (**Figure 2A**). Previous work have showed that tissue-resident CD169^+^ macrophages are positioned in the splenic MZ and reside in the subcapsular sinus of LNs.^7,16^ The strategic anatomical positioning of CD169^+^ macrophages in the spleen and LNs suggest these cells are the first to interact with invading pathogen. Here, we also found that most CD169^+^ macrophages are positive for viral NP antigens (**Figure 2B**). We therefore reasoned that the CD169^+^ macrophages play a central role in mediating SARS-CoV-2 translocation in spleens and LNs, and thus contribute to viral growth and spread.

The presence of viral antigens would activate T cells and B cells in spleen and LNs, and thus lead to Fas and FasL mediated AICD.^17,19^ Although the level of FasL in spleens and LNs is similar in tissues from postmortem and age-match normal healthy controls, the expression of Fas is dramatically higher in virus infected tissue than healthy controls (**Figure 4**). In addition to antigen persistence stimulation, some proinflammatory cytokines like IL-6, TNF-α and IFN-γ can also can mediate cellular apoptosis and necrosis.^20,21^ We focused here on IL-6 because it contributes to host defense against to several kinds of pathogen infections.^21,29^ We also reported that the serum concentration of IL-6 is negative to blood CD4^+^ and CD8^+^ T cells in COVID-19 patients.^14^ Interestingly, exposure to viral Spike protein can trigger *Il6* gene transcription in macrophages *in vitro*, and SARS-CoV-2 infected macrophage also manifested high levels of IL-6 *in vivo* (**Figure 5**).^22,23^ We thereby conjecture that macrophages-derived IL-6 would trigger lymphocyte apoptosis and necrosis, while also enhancing Fas expression. Tocilizumab, a humanized anti-IL-6 receptor antibody, has been developed and approved for the treatment of rheumatoid arthritis (RA).^30,31^ Tocilizumab has also been shown to be effective against cytokine release syndrome resulting from CAR-T cell infusion against B cell acute lymphoblastic leukemia.^32^ Since Tocilizumab has now been approved for use in clinical therapy for COVID-19 patients, whether tocilizumab can restore spleen and LN functions in COVID-19 patients by suppressing IL-6 signaling is an area requiring further investigation.

In conclusion, we have demonstrated that the SARS-CoV-2 virus can directly infect human LNs and spleens and consequently lead to tissue damage and lymphocyte reduction. We also showed that CD169^+^ macrophages play a central role in mediating SARS-CoV-2 translocation. Mechanistically, we observed that viral infected macrophages can promote lymphocyte apoptosis though enhancing IL-6 mediated Fas upregulation.

## Data Availability

For protection of patients' privacy, all data are only provided by authors with anonymous version.

## Authors’ contributions

Yuzhang Wu, and Yongwen Chen were involved in the final development of the project and manuscript preparation; Zilin Yuan, Chenghui Wang and Zeqing Feng analyzed the data; Bo Diao, Yin Liu, Gang Wang, Yinjun Tan and Yueping Liu did H&E staining and immunohistochemistry; Changsong Wang evaluated H&E and immunohistochemistry results; Liang Liu, Rongshuai Wang and Liang Ren provided autopsies and analyzed H&E staining.

## Ethics committee approval

This study was approved by the National Health Commission of China and Ethics Commission of General Hospital of Central Theatre Command and Jinyintan Hospital.

## References

1. Li Q, Guan X, Wu P, et al. Early Transmission Dynamics in Wuhan, China, of Novel Coronavirus-Infected Pneumonia. N Engl J Med 2020.

2. Zhu N, Zhang D, Wang W, et al. A Novel Coronavirus from Patients with Pneumonia in China, 2019. N Engl J Med 2020.

3. Lu R, Zhao X, Li J, et al. Genomic characterisation and epidemiology of 2019 novel coronavirus: implications for virus origins and receptor binding. Lancet 2020.

4. Xu XW, Wu XX, Jiang XG, et al. Clinical findings in a group of patients infected with the 2019 novel coronavirus (SARS-Cov-2) outside of Wuhan, China: retrospective case series. BMJ 2020; 368: m606.

5. World Health Organization. Novel Cornoavirus (COVID-19) Situation, https://experience.arcgis.com/experience/685d0ace521648f8a5beeeee1b9125cd. (Date accessed: March 27, 2020).

6. Aichele P, Zinke J, Grode L, Schwendener RA, Kaufmann SH, Seiler P. Macrophages of the splenic marginal zone are essential for trapping of blood-borne particulate antigen but dispensable for induction of specific T cell responses. J Immunol 2003; 171(3): 1148–55.

7. Mebius RE, Kraal G. Structure and function of the spleen. Nat Rev Immunol 2005; 5(8): 606–16.

8. Junt T, Moseman EA, Iannacone M, et al. Subcapsular sinus macrophages in lymph nodes clear lymph-borne viruses and present them to antiviral B cells. Nature 2007; 450(7166): 110–4.

9. Tamburini BA, Burchill MA, Kedl RM. Antigen capture and archiving by lymphatic endothelial cells following vaccination or viral infection. Nature communications 2014; 5: 3989.

10. van Dinther D, Veninga H, Iborra S, et al. Functional CD169 on Macrophages Mediates Interaction with Dendritic Cells for CD8(+) T Cell Cross-Priming. Cell reports 2018; 22(6): 1484–95.

11. Bernhard CA, Ried C, Kochanek S, Brocker T. CD169+ macrophages are sufficient for priming of CTLs with specificities left out by cross-priming dendritic cells. Proc Natl Acad Sci U S A 2015; 112(17): 5461–6.

12. Backer R, Schwandt T, Greuter M, et al. Effective collaboration between marginal metallophilic macrophages and CD8+ dendritic cells in the generation of cytotoxic T cells. Proc Natl Acad Sci U S A 2010; 107(1): 216–21.

13. Gebhardt T, Mueller SN, Heath WR, Carbone FR. Peripheral tissue surveillance and residency by memory T cells. Trends Immunol 2013; 34(1): 27–32.

14. Diao B, Wang C, Tan Y, et al. Reduction and Functional Exhaustion of T Cells in Patients with Coronavirus Disease 2019 (COVID-19). medRxiv 2020: 2020.02.18.20024364.

15. Diao B, Wang C, Wang R, et al. Human Kidney is a Target for Novel Severe Acute Respiratory Syndrome Coronavirus 2 (SARS-CoV-2) Infection. medRxiv 2020: 2020.03.04.20031120.

16. Moran I, Grootveld AK, Nguyen A, Phan TG. Subcapsular Sinus Macrophages: The Seat of Innate and Adaptive Memory in Murine Lymph Nodes. Trends Immunol 2019; 40(1): 35–48.

17. Strasser A, Jost PJ, Nagata S. The many roles of FAS receptor signaling in the immune system. Immunity 2009; 30(2): 180–92.

18. Alderson MR, Tough TW, Davis-Smith T, et al. Fas ligand mediates activation-induced cell death in human T lymphocytes. J Exp Med 1995; 181(1): 71–7.

19. Krammer PH. CD95’s deadly mission in the immune system. Nature 2000; 407(6805): 789–95.

20. Blaser H, Dostert C, Mak TW, Brenner D. TNF and ROS Crosstalk in Inflammation. Trends Cell Biol 2016; 26(4): 249–61.

21. Tanaka T, Narazaki M, Kishimoto T. IL-6 in inflammation, immunity, and disease. Cold Spring Harb Perspect Biol 2014; 6(10): a016295.

22. Wang W, Ye L, Ye L, et al. Up-regulation of IL-6 and TNF-alpha induced by SARS-coronavirus spike protein in murine macrophages via NF-kappaB pathway. Virus Res 2007; 128(1-2): 1–8.

23. Dosch SF, Mahajan SD, Collins AR. SARS coronavirus spike protein-induced innate immune response occurs via activation of the NF-kappaB pathway in human monocyte macrophages in vitro. Virus Res 2009; 142(1-2): 19–27.

24. Huang C, Wang Y, Li X, et al. Clinical features of patients infected with 2019 novel coronavirus in Wuhan, China. Lancet 2020.

25. Wang D, Hu B, Hu C, et al. Clinical Characteristics of 138 Hospitalized Patients With 2019 Novel Coronavirus-Infected Pneumonia in Wuhan, China. JAMA 2020.

26. Zhang JJ, Dong X, Cao YY, et al. Clinical characteristics of 140 patients infected with SARS-CoV-2 in Wuhan, China. Allergy 2020.

27. Zhan J, Deng R, Tang J, et al. The spleen as a target in severe acute respiratory syndrome. The FASEB Journal 2006; 20(13): 2321–8.

28. Ding Y, Wang H, Shen H, et al. The clinical pathology of severe acute respiratory syndrome (SARS): a report from China. J Pathol 2003; 200(3): 282–9.

29. Jones SA, Jenkins BJ. Recent insights into targeting the IL-6 cytokine family in inflammatory diseases and cancer. Nature Reviews Immunology 2018; 18(12): 773–89.

30. Burmester GR, Rigby WF, van Vollenhoven RF, et al. Tocilizumab in early progressive rheumatoid arthritis: FUNCTION, a randomised controlled trial. Annals of the rheumatic diseases 2016; 75(6): 1081–91.

31. Genovese MC, McKay JD, Nasonov EL, et al. Interleukin-6 receptor inhibition with tocilizumab reduces disease activity in rheumatoid arthritis with inadequate response to disease-modifying antirheumatic drugs: the tocilizumab in combination with traditional disease-modifying antirheumatic drug therapy study. Arthritis Rheum 2008; 58(10): 2968–80.

32. Le RQ, Li L, Yuan W, et al. FDA approval summary: tocilizumab for treatment of chimeric antigen receptor T cell□induced severe or life□threatening cytokine release syndrome. The oncologist 2018; 23(8): 943.

